# Neoadjuvant ablative radiation plus immune therapy favorably remodels the hepatocellular carcinoma tumor microenvironment

**DOI:** 10.1101/2025.11.03.25339412

**Authors:** Zoe Cosner, Zhoubo Guo, Cole Nawrocki, Bidish Patel, Hannah J. Roberts, Kenneth K. Tanabe, Kelsey S. Lau-Min, Elizabeth P. Walsh, Jeffrey W. Clark, Motaz Qadan, Cristina R. Ferrone, Theodore S. Hong, Linda T. Nieman, David T. Ting, Jennifer Y. Wo, Joseph W. Franses

## Abstract

Although immune therapy regimens have significantly improved treatment options for patients with advanced hepatocellular carcinoma (HCC), optimal use of these regimens in earlier disease stages remains poorly defined. We conducted a single-institution, single-arm pilot study (NCT04857684) of neoadjuvant stereotactic body radiation therapy (SBRT) followed by two cycles of atezolizumab plus bevacizumab followed by surgical resection to treat patients with initially resectable HCC (n=8). The primary endpoint was safety as defined by the proportion of patients with grade 3-4 treatment-related adverse events (trAE) by Common Terminology Criteria for Adverse Events (CTCAE) v5.0. Only one patient experienced a grade 3 trAE, 7 of 8 patients proceeded to surgery and all underwent margin-negative (R0) resection; the eight patient did not proceed to surgery due to subsequent disagreement of the resectability of the tumor. One patient experienced a pathologic complete response, and all resected patients were relapse-free as of the data cutoff, with a median follow-up of 16.3 months (2.1–19.9 months). Compared with unmatched, treatment-naïve HCC specimens, post-treatment resection specimens had a significantly higher degree of infiltration by anti-cancer immune infiltrates, including organized peritumoral immune aggregates. Immune infiltration and proximity to tumor cells correlated with pre-operative radiographic response. This study provides a proof-of-concept that combining neoadjuvant SBRT and immune therapy is safe and provides clear rationale for additional prospective clinical studies utilizing this strategy.

## INTRODUCTION

Hepatocellular carcinoma (HCC) is the most common primary liver malignancy and a leading cause of cancer-related mortality worldwide^1^. While surgical resection remains a potentially curative option for early-stage disease, recurrence rates remain unacceptably high, with up to 50% of patients experiencing intrahepatic relapse within three years post-resection^2^. Despite this significant risk, no neoadjuvant or adjuvant therapies have been established to improve long-term survival in resected HCC^3^. In the setting of advanced stage or unresectable disease, combined immune checkpoint inhibitor regimens have emerged as new standards of care for eligible patients, demonstrating improved survival compared to prior monotherapy approaches^4–6^.

Although translating these systemic therapy advances into perioperative strategies remains an ongoing challenge, there have been small, published trials supporting further investigation of preoperative immune therapies in HCC. Ho et al demonstrated that a limited course of neoadjuvant treatment of initially unresectable but liver-confined HCC with cabozantinib plus nivolumab enabled an 80% margin-negative resection rate, with 42% of resected tumors showing major pathological responses and mature immune infiltrates^7^. Kaseb et al showed that a perioperative strategy of 6 weeks of nivolumab with or without ipilimumab prior to resection, resulted in ∼30% of patients with major pathological responses^8^.

Preoperative radiation can reduce the risk of disease recurrence and death even in the localized but high-risk setting of portal vein-invasive, resectable HCC^9,10^. When administered as definitive treatment, stereotactic body radiation therapy (SBRT) has been shown to confer local control rates of greater than 80%^11,12^. Pre-operative radiation may reduce recurrence risk by targeting microscopic residual disease and enhancing local tumor control^13,14^. Beyond local effects, radiation has been shown to modulate the local tumor microenvironment and systemic antitumor immune profiles, potentially acting as an “immune primer”^15,16^. Evidence from other malignancies demonstrates that the combination of radiation and immune checkpoint blockade may potentially result in higher out-of-field response rates compared to immunotherapy alone, supporting the rationale for combining SBRT with immunotherapy in HCC to augment systemic control and reduce recurrence^17,18^.

The combination of SBRT with atezolizumab and bevacizumab has been shown to be safe and effective in treatment of patients with locally advanced HCC^19–21^. We hypothesize that, in the initially resectable HCC disease state, the combination of radiation-primed neoadjuvant immunotherapy (atezolizumab plus bevacizumab) is safe and tolerable and may reduce the likelihood of HCC recurrence compared to up-front surgery alone. In this pilot study, we prospectively evaluated the feasibility and safety of SBRT followed by two cycles of atezolizumab plus bevacizumab prior to surgical resection for initially radiographically resectable HCC. Additionally, we dissected the immune microenvironment of treated resection samples using single cell spatial transcriptomic profiling, finding significant and favorable remodeling of the immune milieu with the abbreviated pre-operative treatment course.

## RESULTS

### Enrolled clinical cohort overview

A schematic overview and CONSORT diagram of trial activities are shown in Fig. 1A-B. Eight patients were enrolled, each of which was initially determined by a surgeon to be eligible for upfront surgical resection. Of note, one patient was deemed upfront resectable by one surgeon and enrolled on study but subsequently transferred care to a different surgeon and was deemed unresectable on re-review of initial baseline assessment. Thus, they did not proceed to surgery after neoadjuvant therapy (Fig 1B). Demographic characteristics are shown in Table 1. Median age at time of enrollment was 71 years (range 65-75 years). Half the patients had viral etiology for development of HCC, with 1 patient (12.5%) having prior Hepatitis B Virus (HBV) and 3 patients (37.5%) having prior Hepatitis C Virus (HCV). The other half of patients had non-viral etiology, with 2 patients (25%) having prior Metabolic Associated Steatohepatitis/Metabolic Dysfunction Associated Steatotic Liver Disease (MASH/MASLD) and 2 patients (25%) having prior alcohol associated liver disease. Most patients had an ECOG performance status of 0 (7, 87.5%), and 1 patient (12.5%) had a performance status of 1. Additionally, most patients (7, 87.5%) had an Alpha Fetoprotein (AFP) level <400, with 1 patient (12.5%) having AFP ≥400. Median Model for End-Stage Liver Disease (MELD) score was 7 (range 6-12). Seven (87.5%) patients had a Childs-Pugh Score of A5, and 1 patient (12.5%) had a score of A6. All patients (8, 100%) had a clinical T stage of T1b. Overall, this cohort was felt to be representative of early-stage HCC patients who undergo resection (Table 1).

**Figure 1.**
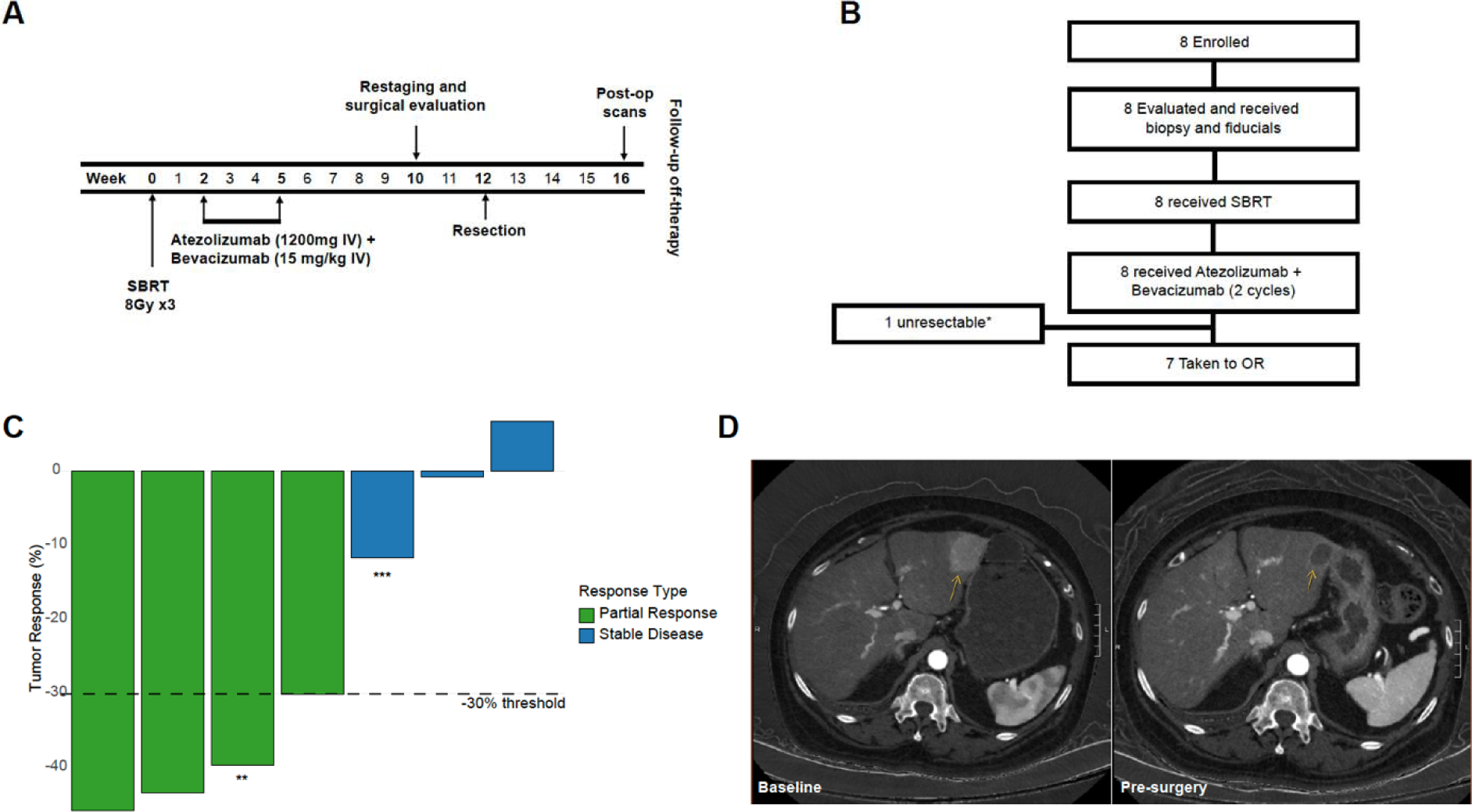
Clinical response characteristics of trial participants. (**A**) Trial evaluation and treatment schema. (**B**) Consort diagram of trial participants’ disposition. * One patient was deemed unresectable due to disagreement after trial enrollment after transfer to another surgeon who disagreed with up front resectability. (**C**) Waterfall plot of radiographic changes, with percent change of post-neoadjuvant treatment compared with pre-treatment baseline. ** Patient with pathologic complete response. *** Patient that did not undergo resection. (**D**) Representative cross-sectional imaging of HCC tumor pre- and post-neoadjuvant treatment.

**Table 1.**
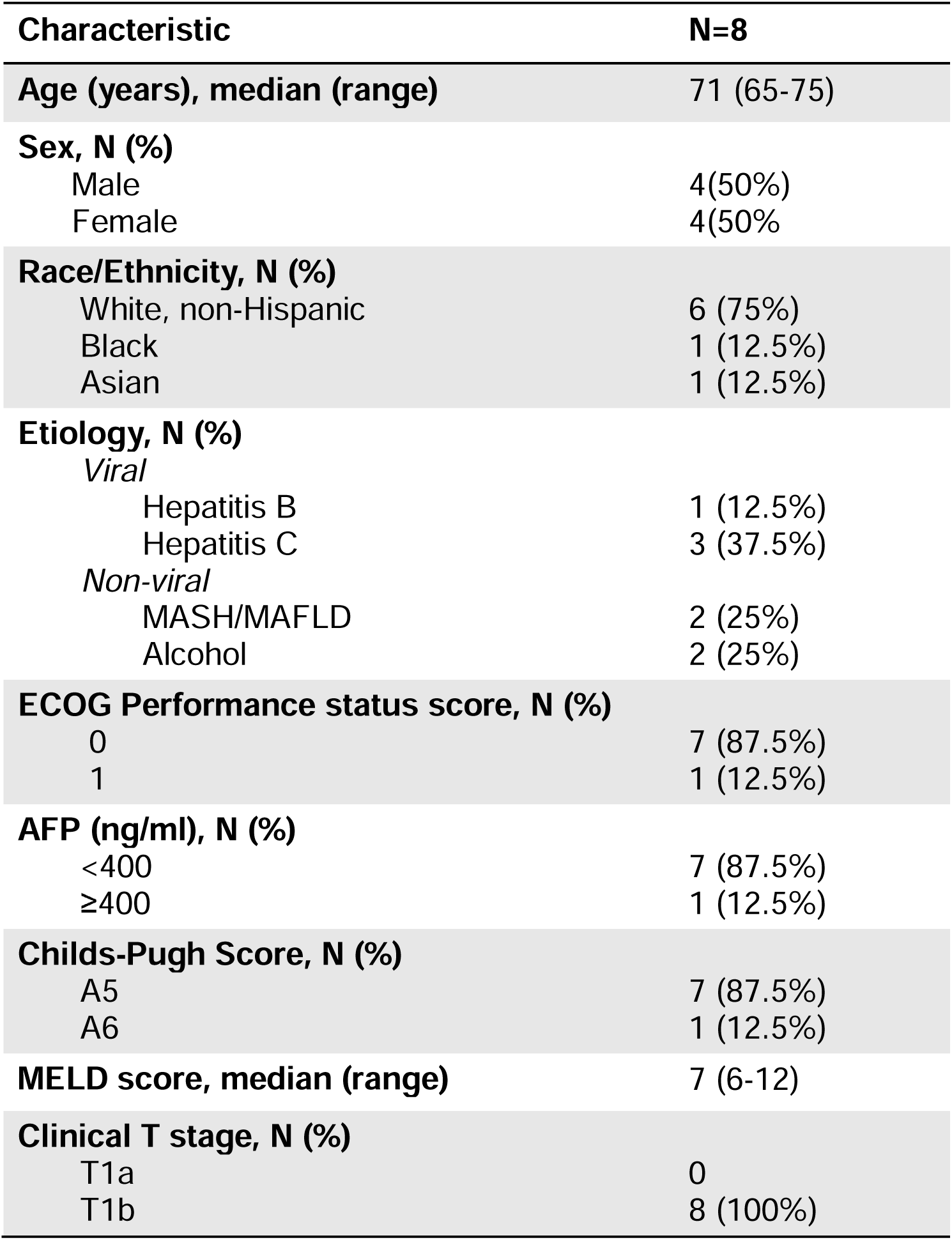
Summary of patient characteristics.

### Safety of the combination regimen

All patients successfully completed SBRT and cycle 1 of atezolizumab/bevacizumab (Table S1). Only one patient experienced a grade 3 treatment related adverse event in the form of hypotension following cycle 1 of atezolizumab/bevacizumab. This patient had the second cycle of atezolizumab/bevacizumab held and proceeded to surgery. The remaining trAEs were not deemed clinically significant (Table S2), and no patients experienced significant post-operative wound healing complications. These data indicate that the combination of neoadjuvant SBRT and atezolizumab/bevacizumab is both feasible, safe, and tolerable.

### Radiographic, pathological, and short-term clinical outcomes predict excellent long-term clinical outcomes

Per protocol, 7 patients had repeated imaging after neoadjuvant therapy and prior to surgery. A protocol deviation occurred leading to omission of the pre-surgery scan for one patient. Of the 7 patients who had imaging (Fig. S1), 4 met the RECIST 1.1 criteria for partial response to therapy, leading to an objective response rate of 57% (Table S3). The remaining 3 patients (43%) had stable disease, with no patients progressing through treatment (Fig. 1C, 1D). Seven of the 8 patients (87.5%) successfully underwent resection following neoadjuvant therapy. One patient (14.3%) achieved a pathological complete response; 3 patients (42.9%) had incomplete necrosis noted. Final pathologic staging included 1 patient (14.2%) with ypT0, 2 patients (28.6%) with ypT1a, 3 patients (42.9%) with ypT1b, and 1 patient (14.2%) with ypT2. Lymphovascular invasion (LVI) was present in 1 patient (14.2%). Each patient who underwent surgery achieved an R0 resection (Table 2). Patients were followed for a median of 16.3 months (range: 2.1-19.9 months) from time of registration to the time of data cutoff. There were no deaths observed, with a 1-year overall survival rate of 100%. Similarly, there were no recurrences observed, yielding a 1-year recurrence-free survival of 100% (Table S3). These results indicate that neo-adjuvant SBRT-primed immunotherapy may provide improved survival benefit and recurrence free survival.

**Table 2.**
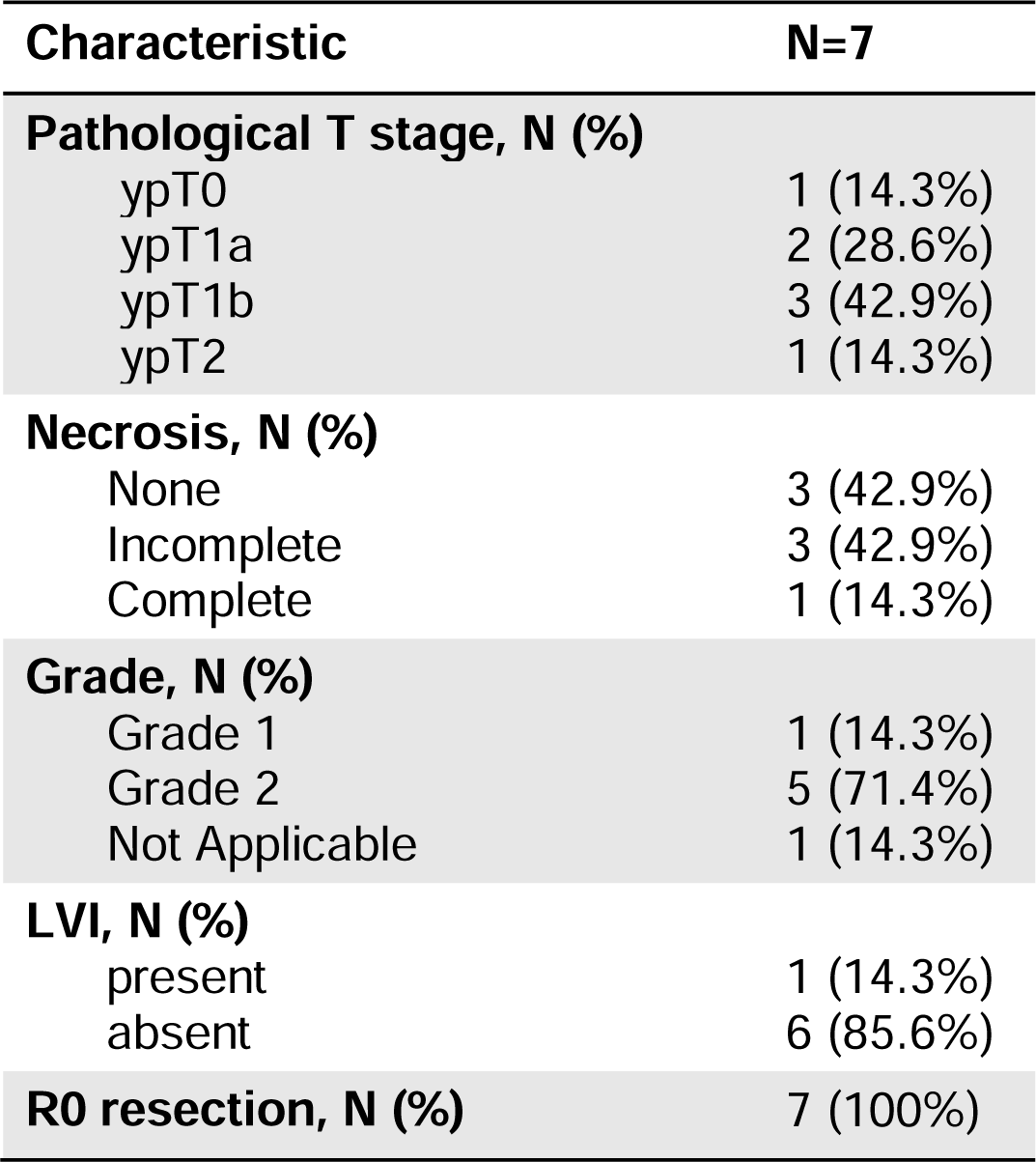
Summary of pathological features in resected tumor specimens.

### Spatial transcriptomic analyses reveal significant immune infiltration after neoadjuvant treatment

To determine whether combined modality neoadjuvant therapy can remodel the immune microenvironment of HCC, we employed spatial molecular imaging (SMI) to a subset of the “post-treatment” (“postTx”) resection specimens, and as a control utilized unmatched SBRT- and systemic therapy-naïve (“TxN”) samples collected from a previous dataset^22^. The experimental workflow is outlined in Fig. 2A. Low-power immunofluorescent images of all included samples illustrating the selection of fields of view (FOVs) are shown in Fig. 2B. We were able to accurately perform cell segmentation (cell membrane boundary identification; Fig. 2C) prior to implementing Uniform Manifold Approximation and Projection (UMAP) dimensionality reduction followed by a combination of supervised and unsupervised cell typing permitted to unambiguously identification of the major cell types present in all the tissue specimens (Fig. 2D, Fig. S2A-C, Table S5). The accuracy of cell type annotations was further validated by comparison of classified cell types with the corresponding immunofluorescence signals (Fig. 2E).

**Figure 2.**
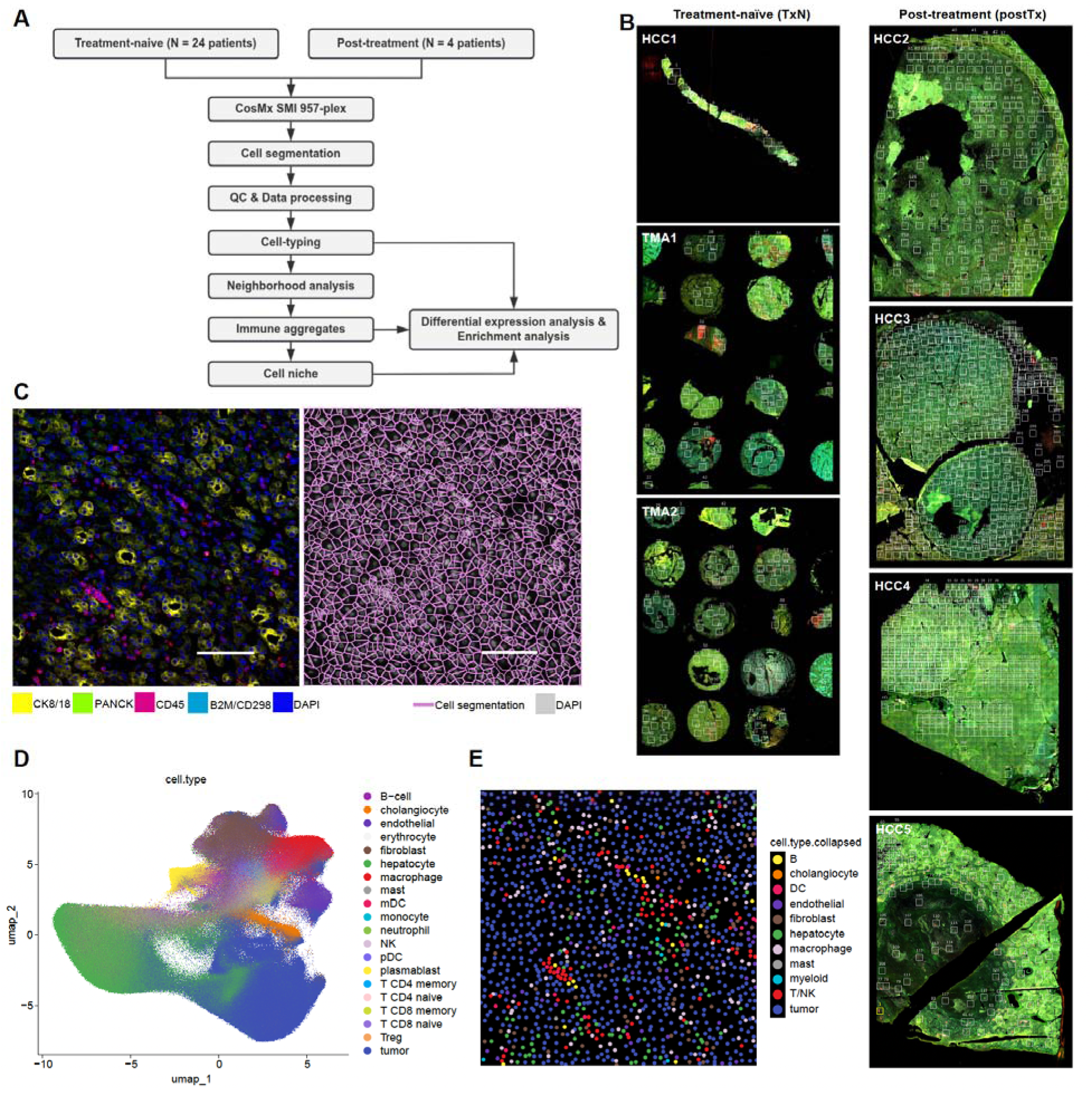
Overview of CosMx spatial transcriptomics data. (A) Schematic overview of single-cell spatial transcriptomics CosMx SMI data acquisition and analysis workflow. (B) Overview of CosMx slides and selection of fields of view (FOVs). Slides HCC1, TMA1, and TMA2 (24 treatment-naïve patient samples) constitute the treatment-naive (TxN) cohort, whereas slides HCC2–HCC5 (four clinical trial patient samples) form the post-treatment (postTx) cohort. Each white box denotes one FOV. (C) Representative fluorescence image (left) and cell segmentation map (right) from the same FOV. Segmentation was based on CK8/18 (yellow), pan-cytokeratin (green), B2M/CD298 (cell membrane; light blue), CD45 (red) and DAPI (nuclei; blue), with magenta lines marking identified cell boundaries. Scale bar represents 100 µm. (D) UMAP visualization of integrated CosMx data, with cells colored by cell type. (E) Spatial map of annotated cell types from the same FOV as in (C), with each point representing an individual cell positioned according to its spatial coordinates.

We identified a total of 1,145,044 cells, including 324,530 from liver FOVs, 683,405 from tumor FOVs, and 137,109 from other FOVs, such as those containing both normal and tumor regions or necrotic areas (Fig. S2D). Within the liver FOVs, the cellular composition consisted of 54.3% hepatocytes, 30.1% immune cells, and 7.62% stromal cells. The tumor FOVs were composed of 54.4% tumor cells, 20% immune cells, and 15.9% stromal cells (Fig. S2E). Across all cells, we detected a median of 163 transcripts and 87 unique genes per cell. The distributions of cell types between the treatment-naïve and post-treatment groups were generally similar (Fig. 3A and S3A), indicating that batch effects had been effectively mitigated.

**Figure 3.**
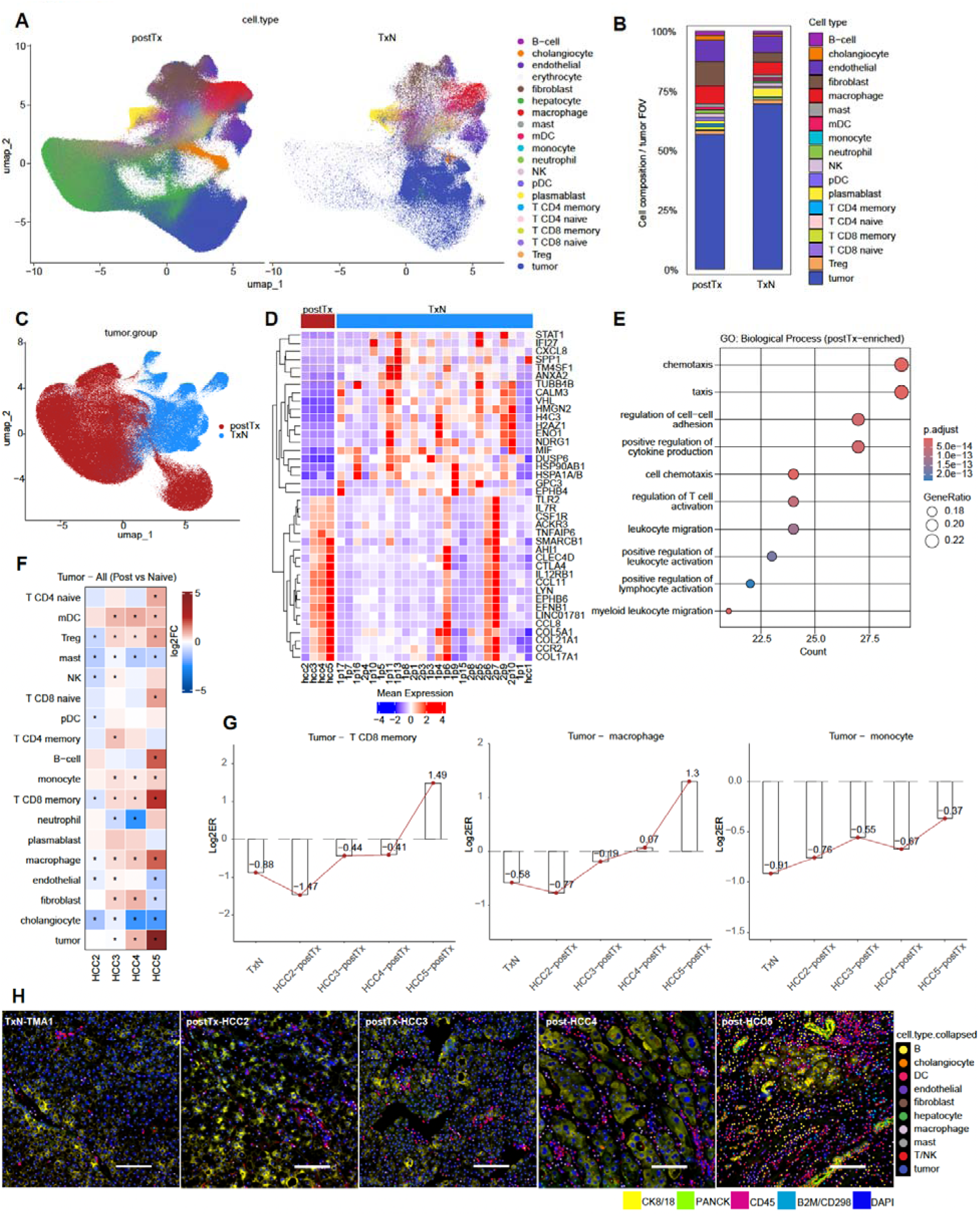
Differential gene expression and spatial analyses between groups. (A) UMAP plots showing all cell types, separated by treatment group (treatment-naïve, TxN, vs. post-treatment, postTx). (B) Proportions of each cell type in tumor FOVs for two groups, with erythrocytes and hepatocytes excluded. (C) UMAP showing tumor cells only, with colors indicating sample group. (D) Heatmap of differential gene expression in tumor cells between treatment-naïve and post-treatment groups, showing the top 20 genes upregulated in each group. (E) Bubble plot showing the top 10 Gene Ontology Biological Process (GO: BP) pathways enriched among genes upregulated in post-treatment tumor cells. (F) FOV-level heatmap of cell-type enrichment around tumor cells. Enrichment ratios compare post-treatment slides with treatment-naive samples, with red indicating greater and blue indicating lower proximity to tumor cells. * indicates p < 0.05. (G) FOV-level log2 enrichment ratios (log2ER) of CD8⁺ memory T cells, macrophages, and monocytes across TxN (combined) and each postTx sample. (H) Representative fluorescence images with cell type annotations from TxN (TMA1) and each postTx slide, highlighting increased immune cell infiltration in HCC3, HCC4, and HCC5. Scale bar represents 100 µm.

However, post-treatment samples contained a lower proportion of tumor cells and a higher proportion of endothelial cells, macrophages, and fibroblasts in tumor FOVs (Fig. 3B and S3B). In addition, we calculated the signature scores of M0, M1, and M2 macrophage phenotypes based on CIBERSORT-derived gene signatures^23^ and compared the differences between two groups. We found that neoadjuvant therapy promoted the polarization of macrophages toward the M1 phenotype rather than M0 or M2 (Fig. S3C). Tumor cells from the two groups showed significant transcriptional differences, as demonstrated by the formation of distinct tumor clusters, which could not be attributed to batch or run effects. (Fig. 3C). The post-treatment tumor cells showed significantly higher expression of immune-related genes (TLR2, CCR2, CCL8, CCL11; Fig. 3D; Fig. S3D; Table S6), and the treatment-naive tumor cells had higher expression of SPP1, TM4SF1, GPC3. Over-representation analysis (ORA) for Gene ontology (GO) biological process (BP) gene sets demonstrated that genes upregulated in post-treatment tumor cells were significantly enriched in pathways related to chemotaxis and cell–cell adhesion (Fig.3E), suggesting heightened crosstalk and recruitment between tumor, stromal, and immune cells. Furthermore, the upregulation of immune-related functional pathways, such as cytokine production, leukocyte activation, and leukocyte migration, indicated robust immune cell activation programs present in the post-treatment tumor cells.

To investigate whether neoadjuvant therapy effectively promotes the recruitment of immune cells into the tumor microenvironment, we applied Delaunay spatial modeling^24^ to define the spatial neighbors of each cell within individual FOVs. Compared with the treatment-naïve group – and particularly in samples HCC3, HCC4, and HCC5 – multiple immune cell types were enriched in proximity to tumor cells within the post-treatment cohort, indicating a tendency for these cells to migrate closer to tumor cells. By contrast, within HCC2, both immune and stromal cells tended to be located further away from tumor cells following neoadjuvant therapy (Fig. 3F), and this correlated with radiographically stable disease after treatment (Fig. S3E) as opposed to radiographic response for the other patients. The enrichment proportions of specific immune cell subtypes – CD8⁺ memory T cells, macrophages, and monocytes (Fig. 3G) – followed a similar pattern, with more significant post treatment-associated enrichment in samples HCC3, HCC4, and HCC5. Representative immunofluorescence images from the treatment-naïve group and each post-treatment sample illustrate the variable degrees of immune cell infiltration across samples (Fig. 3H). Taken together, our results demonstrate that combined modality neoadjuvant therapy promoted a shift of tumor cells toward a more inflammatory phenotype associated with the spatial enrichment of multiple immunologically active immune cell types within the tumor tissue.

### Neoadjuvant therapy promotes immune cell recruitment and the formation of activated immune aggregates

Immune cells often cluster together within tissues to form local immune aggregates that coordinately mediate immune responses. To further characterize the impact of neoadjuvant therapy on the tumor immune microenvironment, we identified immune aggregates in our dataset using both a spatial clustering approach and an unbiased deep learning-based method. First, we applied DBSCAN^25^ spatial clustering on immune and stromal cell coordinate data to detect spatial immune cell aggregates (Fig. 4A). The spatial distribution of immune aggregates across all samples was quite heterogeneous, with significant enrichment of immune cell aggregates at the periphery of the tumors in the post-treatment samples (Fig. S4A), consistent with prior neoadjuvant reports^7^. Post-treatment immune aggregates contained higher proportions of T cells, endothelial cells, and fibroblasts, but lower abundances of plasmablasts, compared with treatment-naïve immune aggregates (Fig.4B). Differential expression analysis between the two groups’ immune aggregates demonstrated that many genes related to immune cell recruitment and activation (CXCL12, IL7R, TNFSF8) were significantly upregulated in post-treatment immune aggregates (Fig. 4C).

**Figure 4.**
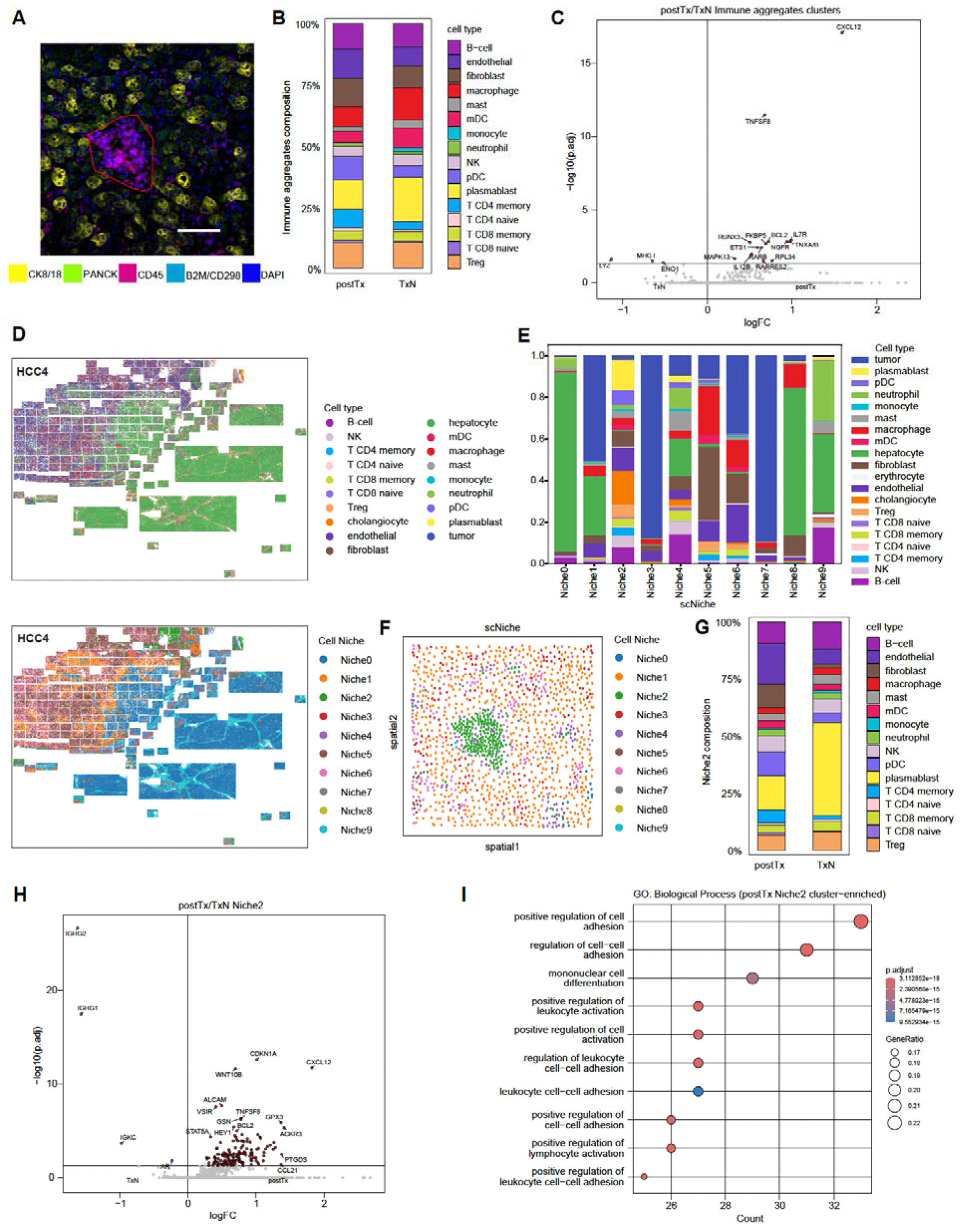
Identification of immune aggregates and cell niches. (A) Representative fluorescence image showing an immune aggregate, with its spatial extent outlined by a convex hull encompassing the constituent immune cells. Scale bar represents 100 µm. (B) Proportions of different cell types within immune aggregates from treatment-naive and post-treatment samples. (C) Differential gene expression in immune aggregate cells between post-treatment and treatment-naive groups in tumor FOVs, visualized as a volcano plot. (D) Identification of ten distinct cell niches across all samples, shown in HCC4 slide area with cell types (top) and cell niches (bottom). (E) Distribution of cell types across different identified niches. (F) Spatial map of a representative FOV (corresponding to A), highlighting Niche 2, which corresponds to immune aggregates identified without prior bias. (G) Cell type proportions in Niche 2 (excluding tumor and cholangiocyte cells) across treatment-naive and post-treatment tumor FOVs. (H) Differential gene expression in Niche 2 cells (excluding tumor and cholangiocyte cells) between post-treatment and treatment-naive groups in tumor FOVs, visualized as a volcano plot. (I) Enriched Gene Ontology (GO) Biological Process terms for genes upregulated in Niche2 of post-treatment samples.

Next, we sought to use unbiased methods to examine immune niches in the tumor microenvironment. We utilized a recently reported graph neural network-based approach, scNiche^26^, that integrates the molecular profile of cells’ spatial neighbors and the composition of neighboring cell types to define cell neighborhoods. We used this algorithm to identify 10 distinct cellular niches (Fig. 4D, Fig. S5A) across all treatment-naïve and post-treatment samples (Fig. S5B). Treatment-naïve samples – including HCC1 and all samples from TMA1 and TMA2 – exhibited a similar niche composition, predominantly composed of Niche7, Niche6, and Niche2, whereas post-treatment samples displayed a more heterogeneous niche distribution (Fig. S5C). Among these, Niche2 was characterized by a cellular composition dominated by various immune and stromal cell types, with only a minor fraction of tumor cells (Fig. 4E, Fig. S5D). Morphologically, cells in Niche2 formed visually discernible aggregates (Fig. 4F), suggesting that Niche2 may represent a structure analogous to immune aggregates (Fig. S6A). When tumor cells and cholangiocytes were excluded, the cellular composition of Niche2 within tumor FOVs from treatment-naïve and post-treatment groups resembled that of the immune aggregates identified in our spatial clustering analysis. Specifically, post-treatment samples contained higher proportions of CD4⁺ T cells, endothelial cells, and fibroblasts, and dendritic cells, whereas B cells and plasmablasts were enriched in treatment-naïve samples (Fig. 4G). Furthermore, the post-treatment Niche2 microenvironment exhibited increased expression of chemokines (*CXCL12*, *CCL21*) and *TNFSF8*, supporting the notion that neoadjuvant therapy promotes the recruitment and migration of immune cells into intratumoral multicellular immune aggregates (Fig. 4H). ORA for GO: BP gene sets further revealed enrichment of processes such as adhesion, lymphocyte activation (Fig. 4I).

Taken together, our findings suggest that neoadjuvant of SBRT plus two infusions of atezolizumab and bevacizumab results in a significant infiltration into and organization of immune aggregates within the tumor milieu. The presence of these maturing aggregates correlates with pre-operative radiographic response, pathologic tumor necrosis, and recurrence-free survival.

## DISCUSSION

In summary, our prospective pilot study was the first to combine neoadjuvant SBRT with combination immune checkpoint inhibitor therapy for patients with initially resectable HCC. We showed that the treatment algorithm was tolerable and did not lead to loss of opportunity for curative-intent, margin-negative resection. Further, we observed a 100% relapse-free survival rate, with a median follow-up of 16 months, for the seven out of eight participants who underwent curative-intent resection; the eighth patient had a second surgical opinion who disagreed with the resectability and hence did not proceed to resection. Notably, this is a higher percentage than expected for surgical resection alone^27^. These findings were correlated with tumor necrosis in four of the seven resected patients and a pathological complete response in one. Our study’s clinical results are consistent with prior published neoadjuvant immune therapy studies without radiation, in which radiographic response, tumor necrosis, relapse-free survival, and overall survival are correlated with one another^7,8^.

Although small, prospective neoadjuvant immune therapy studies in HCC seem to confer promising safety and efficacy endpoints, adjuvant studies have yet to show a clinical benefit. The initially published analysis of the phase 3, randomized IMbrave050 trial^28^ showed an early improvement in relapse-free (but not overall) survival for atezolizumab plus bevacizumab compared with placebo after surgical resection or ablation, but even this benefit was lost with further follow up^29^. This discrepancy in potential efficacy between purely adjuvant immune therapy – in which only microscopic residual disease may be present – and regimens incorporating neoadjuvant therapy – in which bulk tumor tissue is present during immune therapy treatment – is consistent with other tumor types. For example, perioperative pembrolizumab was shown to be superior to adjuvant-only administration for melanoma^30^, and a meta-analysis of multiple non-small cell lung carcinoma trials showed that the addition of adjuvant immune therapy to neoadjuvant immune therapy does not improve clinical outcomes^31^. Therefore, both immune checkpoint inhibitors and certain schedules and doses of radiation therapy^32^ may require tumor bulk to effectively educate or augment anti-tumor immunity.

Our single-cell spatial transcriptomic analysis revealed that high expression of SPP1^33^, TM4SF1^34^, and GPC3^35^ is characteristic of treatment-naive tumor cells, indicating an aggressive and immunosuppressive phenotype. In our study, the TME was reprogrammed by neoadjuvant therapy toward an anti-cancer inflammatory state by combination therapy, especially in samples HCC3–5. Specifically, neoadjuvant treatment induced upregulation of CCR2, CCL8, and CCL11, key CC chemokines and receptors, facilitating immune cell infiltration into the tumor. This is consistent with previous reports showing enhanced intratumoral macrophage infiltration following immunotherapy^36^. Our data also revealed polarization of the recruited macrophages toward the M1 phenotype, which often correlates with antitumor immunity in HCC and other tumor types^37^. Further, upregulation of TLR2 in tumor cells suggested that neoadjuvant therapy promotes T cell immune activation, similar to the work reported by Li et al., demonstrating that TLR2 activation suppresses IL8 expression, thereby reducing myeloid derived suppressor cell (MDSC) infiltration and enhancing the antitumor function of activated CD8⁺ T cells^38^.

Finally, unlike droplet-based single-cell transcriptomics approaches, which sacrifice spatial data during tissue dissociation, our transcriptomic strategy preserved spatial information for analysis. This strategy enabled identification of significant immune cell organization and cancer-immune interactions compared with unmatched systemic therapy-naïve tumors. The induced immune aggregates identified in our study tended to appear at the tumor periphery, a homologous finding to those of other neoadjuvant immunotherapy studies^39^. Further, the composition of our immune aggregates strongly resembles nascent “tertiary lymphoid structures” (TLSs), which tend to correlate with good clinical outcomes in HCC^40^ and other^41^ tumor types. As in Shu et al’s study of the TME in resected HCC tissue after exposure to neoadjuvant immune checkpoint inhibitor regimens^40^, our combined modality treatment regimen correlated with a significant enrichment of the number and complexity of immune aggregates (including lymphoid, myeloid, and endothelial components) within HCC tumors. This immune organization may be driven by certain chemokines identified in our study, including CXCL12 and CCL21, which have been reported to induce the formation of TLSs^42,43^. Notably, VEGF-C or tobacco exposure can activate the CCL21/CCR7 axis, promoting TLS formation, enhancing antitumor immunity, and improving the efficacy of immunotherapy^44,45^.

Although our pilot study had several significant strengths as noted above, there were several limitations in the study design and analysis. First, the small sample size and non-randomized single institution trial design did not allow for direct comparisons with other neoadjuvant strategies (including dose/schedule of the atezolizumab plus bevacizumab regimen or SBRT) or with up front resection. Second, although the single cell spatial transcriptomic approach is unique, the panel-based transcriptomic approach of ∼1,000 genes for analysis necessarily led to omission of ∼95% of the coding transcriptome. Third, the spatial transcriptomic analysis did not capture matched pre- and post-treatment specimens from every participant, necessitating the use of an unmatched treatment-naïve cohort.

In summary, our study is the first to demonstrate the feasibility and safety of neoadjuvant SBRT plus two cycles of atezolizumab plus bevacizumab for the treatment of patients with initially resectable HCC. The combined-modality neoadjuvant therapy led to infiltration and organization of multicellular anti-cancer immune aggregates, which correlated with positive radiographic and pathologic response outcomes and excellent relapse-free survival. This study provides motivation for subsequent randomized studies to evaluate interventions that can augment effective antitumor immune trafficking prior to curative-intent resection for HCC, including the addition of additional agents to stimulate the critical signaling nodes associated with anticancer immune niches.

## METHODS

### Patient Cohort and Eligibility Criteria

Between June 2021 and January 2024, we enrolled 8 eligible patients with treatment-naïve HCC at the Massachusetts General Hospital Cancer Center. The initial participant goal was 20 participants, but ultimately the trial was halted early due to slower-than-anticipated accrual. Eligible patients were 18 years or older with Eastern Cooperative Oncology Group (EGOG) Performance Status (PS) of 0 or 1, Child-Pugh A liver function, no evidence of significant portal hypertension, no major medical contraindication to surgery or general anesthesia, and treatment-naïve hepatocellular carcinoma (HCC). The tumor must have been localized to the liver without radiographic evidence of macrovascular invasion, be deemed surgically resectable, and have measurable disease on CT scan or MRI, defined as at least one lesion that can be accurately measured in at least one dimension as ≥10 mm. For patients with active HBV, patients remained eligible if they were well controlled on anti-HBV treatment at least 14 days prior to treatment initiation, and through entirety of treatment. Patients with HCV remained eligible if infection was resolved or chronic. Additionally, participants must have adequate organ and marrow function as defined by leukocytes ≥ 3000/μL, absolute neutrophil count ≥1500/μL, platelets ≥100,000/μL, total bilirubin ≤2x institutional upper limit of normal (ULN), AST (SGOT) and ALT (SGPT) ≤3 x institutional ULN, and creatinine ≤1.5 x institutional ULN or estimated glomerular filtration rate (eGFR) ≥50 ml/min/1.73m^2^. Patients were excluded if they had known fibrolamellar, sarcomatoid, mixed/biphenotypic or cholangiocarcinoma-HCC, radiographic evidence of metastasis to lymph nodes or other extra-hepatic sites, coinfection with HBV and HCV, history of hepatic encephalopathy or moderate to severe ascites, significant bleeding risk or vasculopathy, prior locoregional treatment, or grade ≥2 proteinuria.

### Study Oversight and Drug Supply

All patients provided written informed consent prior to enrollment. The trial was registered under ClinicalTrials.gov as NCT04857684. The protocol underwent review and was approved by Institutional Review Board at the Dana-Farber Harvard Cancer Center (DFHCC). Atezolizumab and bevacizumab were supplied by Genentech.

### Study Design and Follow-Up

This single-center, open-label, single-arm pilot study was conducted at Massachusetts General Hospital (MGH). The study initially designed to include 20 patients but stopped early with 8 patients due to slower-than-anticipated accrual. Eligible patients were enrolled and underwent pre-treatment biopsy with fiducial marker placement followed by radiation planning/mapping once histologic diagnosis was confirmed. Patients received 3 fractions of 8 Gy SBRT to the primary tumor site, administered on an every-other-day basis, followed by two 3-week cycles of atezolizumab (1200 mg IV on day 1) plus bevacizumab (15 mg/kg IV on day 1). After completion of systemic therapy, patients underwent radiographic restaging and surgical re-evaluation. If the patient’s disease remained resectable, patient underwent surgical resection.

Post-surgery scans and continued every 12 weeks until disease recurrence or 2 years post-surgery. Tumor assessments were performed at baseline and prior to surgical re-evaluation. Baseline evaluations were conducted within 1 week prior to start of protocol therapy with baseline scans ≤6 weeks prior to start of therapy. Triple-phase contrast-enhanced abdominal MRI or CT abdomen/pelvis were obtained at each time point. Adverse effects were assessed using National Cancer Institute Common Terminology Criteria for Adverse Events v5.0 (CTCAE v5.0).

The primary study endpoints were safety and tolerability, defined by the proportion of patients with grade 3-4 treatment-related adverse events as assessed by CTCAE v5.0. Secondary endpoints included objective response rate (ORR) after SBRT and atezolizumab and bevacizumab per RECIST 1.1, proportion or patients whose disease remained resectable after neoadjuvant treatment, proportion of patients who undergo a microscopic margin-negative (R0) resection, proportion of patients whose cancer is resected in whom there is a pathologic complete response (CR) of the tumor, overall survival (OS), and recurrence-free survival (RFS) after resection.

### Statistics for clinical data

Relapse-free survival (RFS) was defined as the time between trial registration to local or distant relapse or last on-study assessment. Overall survival was defined as the time between registration to death or status at last on-study assessment of survival. Objective response rate (ORR) was calculated per Response Criteria in Solid Tumors version 1.1 (RECIST 1.1), with clinical partial response designated as tumor diameter decrease of ≥30% and no new lesions observed. Kaplan-Meier estimates characterized time-to-event endpoints. The target accrual was 20 participants, but the trial was halted early, after 8 participants were enrolled, due to slow accrual.

### Spatial transcriptomics cohorts, sample processing and data acquisition

Tissue sections from four patients enrolled in the clinical trial were included and designated as HCC1–HCC5. Among these, HCC2–HCC5 were obtained after neoadjuvant therapy, whereas HCC1 and HCC5 originated from the same patient, with HCC1 representing a pre-treatment needle biopsy specimen. Following neoadjuvant therapy, patients corresponding to samples HCC4 and HCC5 exhibited >30% tumor reduction on imaging, achieving partial response, while the patient corresponding to HCC2 achieved stable disease. Post-treatment imaging for the patient corresponding to sample HCC3 was unavailable. In addition, we included, as a comparison, a treatment-naïve HCC cohort using the same CosMx SMI 957-plex probe set^22^. The treatment-naïve cohort comprised two TMAs (TMA1 and TMA2) containing a total of 23 HCC samples. Consequently, the final study cohort consisted of 24 treatment-naïve samples, 23 from TMAs and one from the HCC1 needle biopsy, and 4 post-treatment samples from clinical trial patients.

The protocol for sample processing and data acquisition has been described in detail in our previous study^22^. Briefly, 5-µm formalin-fixed, paraffin-embedded (FFPE) tissue sections were retrieved from the pathology archives. FFPE sections were baked, deparaffinized, and subjected to proteinase K digestion and heat-induced epitope retrieval using a Leica Bond Rx system. After fiducial marker incubation, fixation, and blocking, probe hybridization (957 gene targets) was performed, followed by stringent washes and blocking. The 957-gene panel is provided in Table S4. The prepared slides were assembled into a flow cell and imaged on the NanoString CosMx SMI platform for imaging.

### Spatial transcriptomics data processing and quality control

The CosMx data processing and analysis pipeline used in this study was adapted from the workflow described by Coley et al^22^, and all processing and analyses were conducted using R 4.4.1. Each dataset was subjected to independent quality-control and processing steps. We examined and selected FOVs (fields of view) from all slides, and their tissue types were annotated based on pathological review. Cell segmentation was performed using the Cellpose algorithm^46^ on fluorescence images stained for pan-cytokeratin (PANCK), keratin 8/18 (CK8/18), CD45, B2M/CD298, and DAPI. Low-quality FOVs were removed using NanoString’s custom runFOVQC function. Low-quality cells were filtered if they had fewer than 20 or more than 2000 total RNA counts, a cell area smaller than 25 µm² or larger than 1000 µm², more than 15% of counts coming from negative control probes, or a complexity score – defined as the log_10_ ratio of total counts to the number of positive genes – greater than 1. To preserve the spatial structure of the tissue, FOVs were excluded entirely if more than 10% of their cells failed any of these quality metrics.

Next, the data was total counts normalized and log-transformed, and individual datasets were merged into a single Seurat object. The logged and normalized data was then scaled before principal component analysis (PCA) was performed. The 50 principal components that accounted for the most variance were retained. To correct for batch effects between sample types, Harmony integration^47^ was applied using dataset identity as the grouping variable. The first 25 Harmony components were then used to generate a unified low-dimensional embedding via UMAP for visualization.

### Cell-typing and validation

To identify tumor and hepatocyte populations, we first applied unsupervised clustering using InSituType, developed by NanoString (Danaher et al., bioRxiv 2022), across all cells. Clusters were annotated and validated based on the expression of cluster-specific genes and by reviewing immunofluorescence images from each FOV. Supervised cell-type annotation was then performed on the remaining cells using InSituType, guided by liver^48^ and immuno-oncology reference profiles. During clustering with the immuno-oncology reference, genes suspected to be affected by spatial contamination were excluded to reduce background noise. To further validate cell-type assignments, we used a pseudo-bulk differential expression approach. Gene-level analysis was conducted using the presto-GLMM R package, which fits a Poisson generalized linear mixed model (GLMM) for each gene. The model estimated random effects for cluster, patient, and their interaction, including total UMI counts as an offset. For each cluster, a gene’s differential expression was assessed by comparing it to all others using linear contrasts, and significance was determined via Wald tests with FDR correction. Genes with a log fold change (logFC) greater than 1.5 and an adjusted p-value less than 0.05 were identified as marker genes. The marker genes we identified in this manner confirmed the accuracy of our clustering and cell-type annotation results. For better visualization, in some plots, cell types were collapsed as follows: “B cells” included B-cell and plasmablast; “dendritic cells” included mDC and pDC; “T and NK subsets” included NK, T CD4 memory, T CD4 naive, T CD8 memory, T CD8 naive, and Treg; “myeloid” included monocytes and neutrophils. Cholangiocytes, endothelial cells, fibroblasts, hepatocytes, tumor cells, and erythrocytes were kept as originally annotated.

### Differential Expression Analysis and Gene Set Enrichment Analysis

We used the NEBULA^49^ package to model gene expression in tumor, endothelial, macrophage, fibroblast, Treg, CD4⁺ memory T, CD8⁺ memory T, B, and plasmablast cells using a negative binomial mixed-effects model. Differential expression between cells from the post-treatment and treatment-naïve groups was assessed separately for each cell type using a Wald test, with p-values adjusted for multiple testing using the false discovery rate (FDR). Genes likely affected by spatial contamination were excluded from the analysis. Genes significantly upregulated in the post-treatment group were extracted for downstream enrichment analysis. ORA for GO:BP gene sets was performed using the clusterProfiler R package^50^. The top 10 enriched biological process pathways, ranked by the number of enriched genes, were visualized using a bubble plot.

### Neighborhood Enrichment Analysis

Neighborhood enrichment analysis was performed following the approach described by Varrone et al^51^. For each FOV, we constructed a Delaunay tessellation to define direct spatial neighbors. Cells were considered neighbors if they were connected in the graph, located within the same FOV, and within 100 µm of each other. To assess spatial organization, we analyzed each cell type separately. For each type, index cells were defined, and the proportions of neighboring cell types were calculated and compared to expected proportions derived from the global edge distribution. A small pseudo-count was added, and log_2_(observed/expected) was computed to obtain the log-normalized enrichment score (log2ER).

To compare global cell colocalization patterns between the post-treatment and treatment-naïve groups, neighborhood graphs were constructed separately for all FOVs from each group. Log2ER values were calculated, and group-level differences were assessed via permutation testing. Specifically, FOV group labels were shuffled 1,000 times, and the resulting null distribution of log2ER differences was used to compute empirical p-values.

### Identification of immune aggregates and cell niches

To identify immune aggregates, we performed spatial clustering using the DBSCAN^25^ algorithm based on cell coordinates and cell type annotations from the CosMx dataset. Specifically, we extracted a subset of cells including T cells, B cells, macrophages, plasmablasts, dendritic cells (DCs), mast, natural killer (NK) cells, monocytes, and neutrophils, fibroblasts, and endothelial cells. Cells were classified as part of a potential immune aggregate cluster if they were located within a 20 µm radius of at least 9 other cells from the specified types (i.e., forming a cluster of ≥10 immune-related cells), and if the total number of cells within the immune aggregate cluster was ≥30. Differential gene expression analysis between immune aggregates in postTx and TxN groups was performed using the NEBULA^49^ package.

To identify spatial niches, we applied the scNiche framework^26^ implemented in Python 3.9.23. This model integrates three key features: each cell’s molecular profile, the molecular profile of its spatial neighbors, and the composition of neighboring cell types. Harmony-reduced dimensions were used as input. Spatial neighbors and spatial expression were computed using K-nearest neighbors (KNN) with k = 10. The model was trained in batches (100 batches) with a learning rate of 0.01 over 100 epochs. After training, unsupervised clustering with k-means was used to identify distinct cell niches. Downstream visualization and niche-level comparisons were performed using Scanpy^52^, NEBULA^49^ package, and clusterProfiler package^53^.

### Data sharing

All data and code used in this study will be made available upon request.

## Data Availability

All data and code used in this study will be made available upon request.

## ACKNOWLEDGMENTS

The authors would like to thank the clinical trial group at Massachusetts General Hospital for critical assistance with clinical trial data monitoring and abstraction. The authors also thank Genentech for supporting the clinical trial operations.

## ETHICS STATEMENT

Ethics Committee/IRB of Massachusetts General Hospital gave ethical approval for this work.

