## Supplemental Table 1-3 and Figure S1-S6 for "Neoadjuvant ablative radiation plus immune therapy favorably remodels the hepatocellular carcinoma tumor microenvironment"

**SUPPLEMENTARY TABLES**

| **Stage of treatment** | **Delay in treatment N (%)** | **Treatment Omitted** |
| --- | --- | --- |
| **SBRT** | **1 (12.5%)** | **0** |
| **Cycle 1 Atezo/Bev** | **1 (12.5%)** | **0** |
| **Cycle 2 Atezo/Bev** | **1 (12.5%)** | **2 (25%)** |

**Table S1. Summary of treatment delays and omissions.**

| **Stage of treatment** | **Adverse Event** | **Grade (Patient number)** |
| --- | --- | --- |
| Baseline | Increased Lipase  Decreased Blood counts  Hyperglycemia | Grade 4 (1)  Grade 2 (1), Grade 3 (1)  Grade 2 (1) |
| After SBRT | Fatigue | Grade 2 (1) |
| After Cycle 1 Atezo/Bev | Hypotension  Hyperglycemia  Decreased blood counts  GI side effect | Grade 3 (1*)  Grade 1 (1*)  Grade 2 (1)  Grade 1 (1) |
| After Cycle 2 Atezo/Bev | Increased lipase  Hyperglycemia  Hypertension  Fatigue | Grade 1 (1)  Grade 1 (1)  Grade 1 (1)  Grade 1 (1) |
| Unrelated to treatment | Constipation  Increased CPK  Leg pain | Grade 1 (1)  Grade 1 (1)  Grade 2 (1) |

**Table S2. Summary of adverse events in study participants.**

| **Statistic** | **value** |
| --- | --- |
| **Proceeded to surgery, N (%)** | **7 (87.5%)** |
| **pCR, N (%)** | **1 (14.3%)** |
| **ORR** | **57%** |
| **Follow up, Median (range)** | **16.3 months (2.1 -19.9 months)** |
| **OS** | **100%** |
| **Recurrence-Free Survival** | **100%** |

**Table S3. Summary of surgical and clinical outcomes.**

**Table S4. List of genes included in the CosMx SMI panel.**

**Table S5. Highly expressed marker genes for each annotated cell type.**

**Table S6. Differential expression analysis results comparing post-treatment (postTx) versus treatment-naïve (TxN) samples for each cell type.**

***** Tables S4–S6 are provided as three separate spreadsheet files. *****

**SUPPLEMENTARY FIGURES**

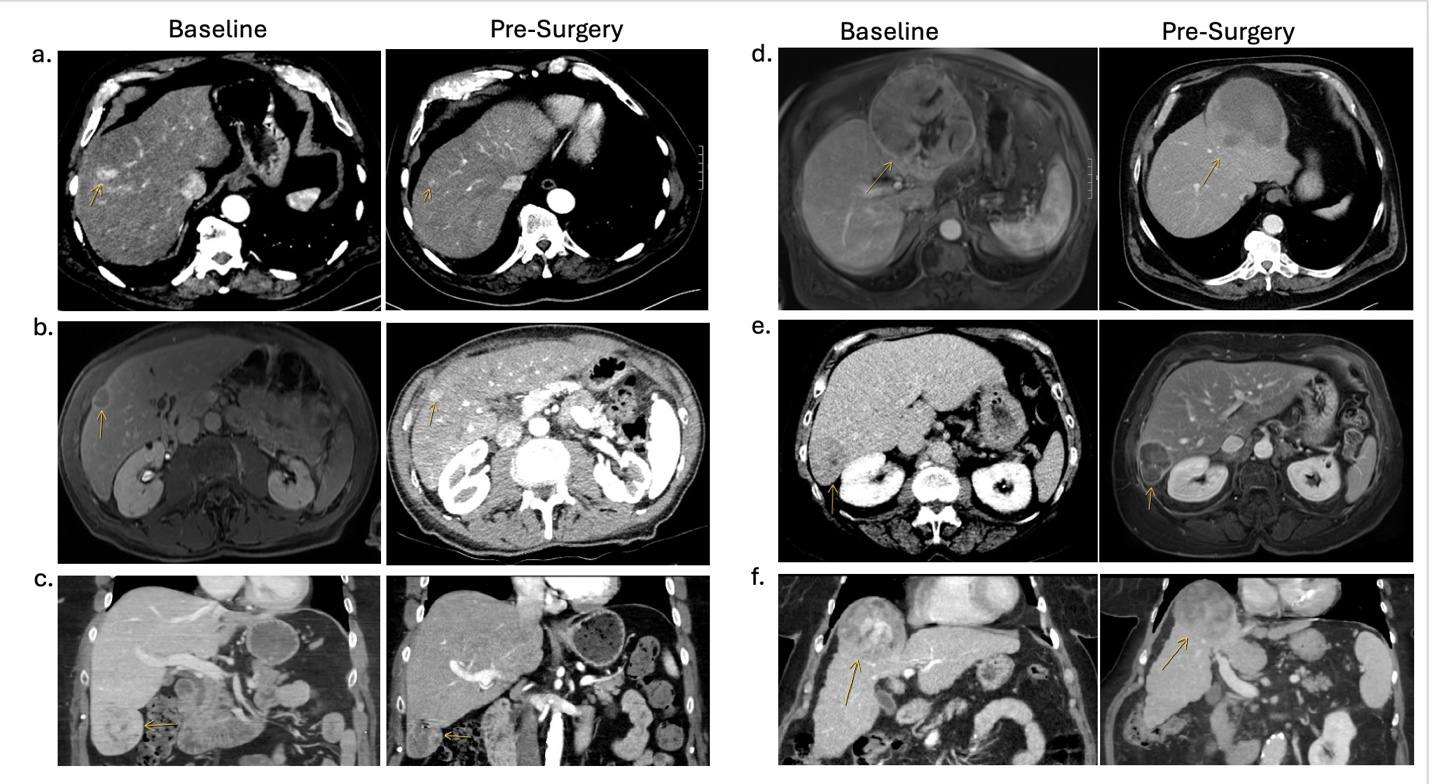

**Figure S1.** Cross-sectional imaging showing baseline and pre-surgery CT scans of remaining patients that underwent both sets of imaging. (**A-C**) Patients had partial response per RECIST 1.1 criteria. (**D-F**) Patients had stable disease per RECIST 1.1 criteria.

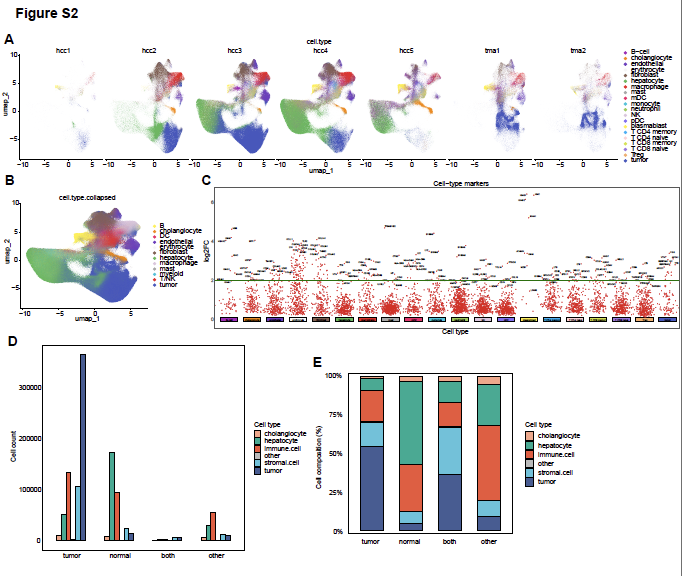

**Fig. S2. Cell type annotation and data information.** (**A**) UMAP plots of all cells, colored by cell type, divided by individual sample. (**B**) UMAP of cells colored by collapsed cell type. (**C**) Manhattan plot showing genes significantly upregulated in each annotated cell type identified by differential expression analysis. Absolute numbers (**D**) and proportions (**E**) of tumor cells, hepatocytes, immune cells, stromal cells, and cholangiocytes across different FOV classes.

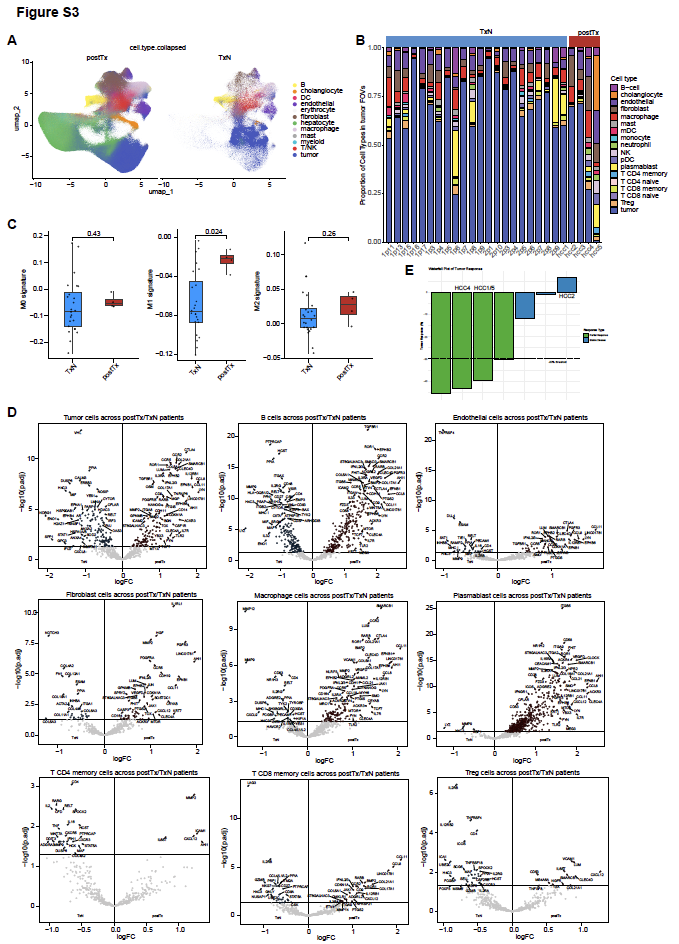
 **Fig. S3.** **Cell type composition and comparative analyses.** (**A**) UMAP of cells colored by collapsed cell type and split by slide. (**B**) Proportions of cell types across individual samples. (**C**) Comparison of macrophage subtype scores (M0, M1, and M2) between postTx and TxN groups. (**D**) Differential gene expression between postTx and TxN samples in macrophages, tumor cells, endothelial cells, fibroblasts, T cells, B cells, and plasmablasts, visualized as volcano plots. (**E**). Treatment responses of patients corresponding to the single cell spatial transcriptomic samples.

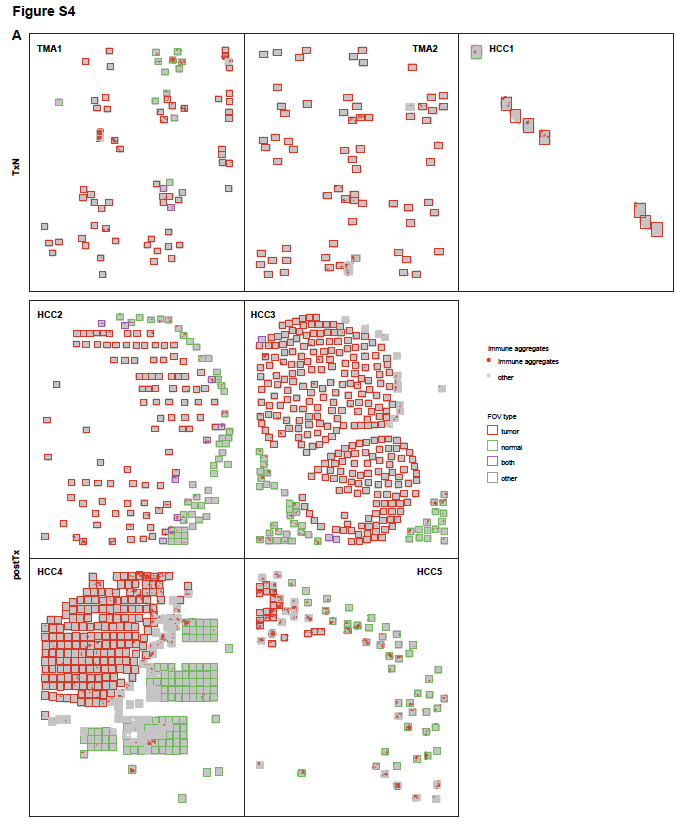

**Fig. S4.** **Spatial localization of immune aggregates.** Spatial distribution of immune aggregates in TMA1, TMA2, HCC1, HCC2, HCC3, HCC4, and HCC5 slides. Red dots indicate cells within immune aggregates; gray dots represent other cells. Red boxes indicate tumor FOVs, green boxes indicate normal liver FOVs, purple boxes represent both FOVs (containing both normal liver regions and malignant cell regions), and gray boxes indicate other FOVs.

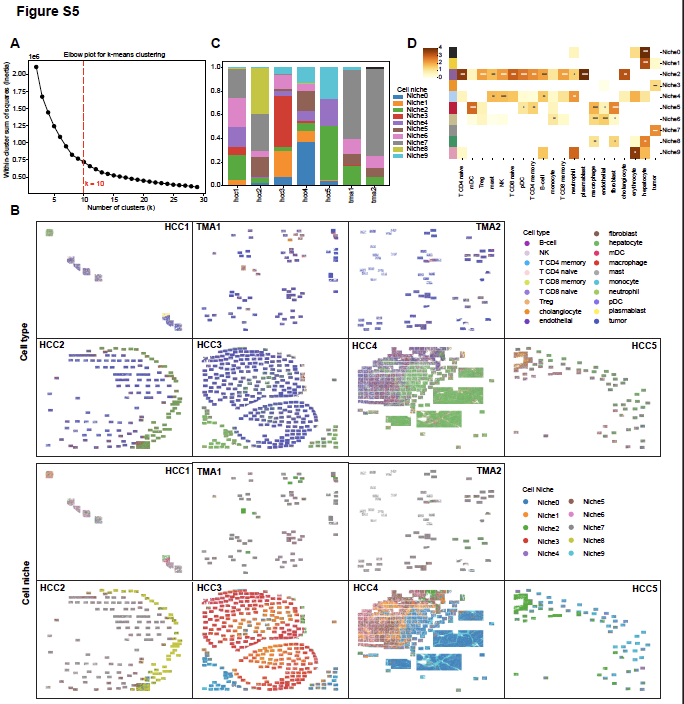

**Fig. S5.** **Identification and characterization of cell niches.** (**A**) Elbow plot used to determine the optimal number of cell niches. (**B**) Distribution of cell types and identified cell niches across all slides. (**C**) Niche proportions across all samples. (**D**) Heatmap shows the enrichment levels of each cell type within the respective cell niches, * indicates p < 0.05, * indicates p < 0.01, * indicates p < 0.001.

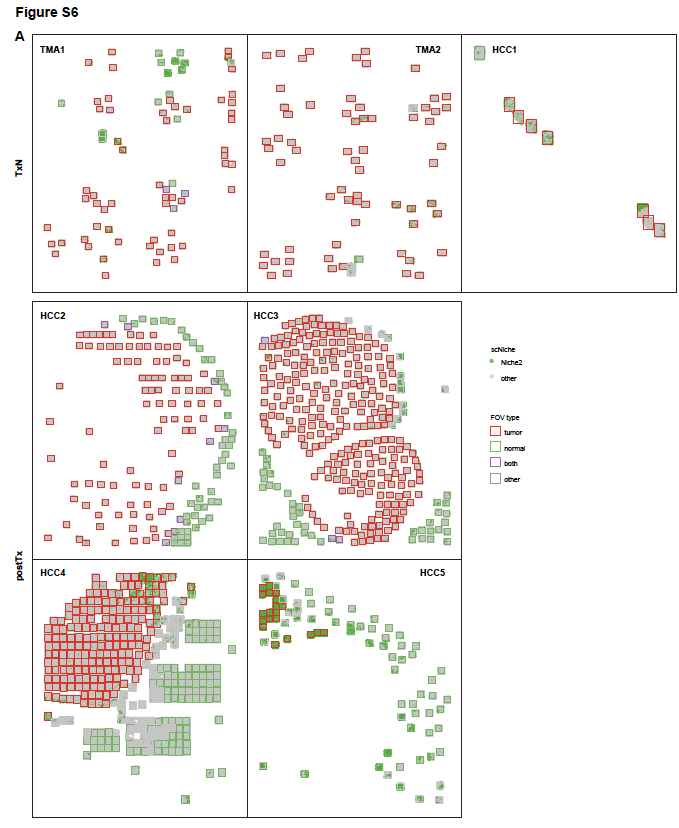

**Fig. S6.** **Spatial localization of Niche2 cells.** Spatial distribution of Niche2 cells in TMA1, TMA2, HCC1, HCC2, HCC3, HCC4, and HCC5 slides. Green dots indicate cells belong to Niche2; gray dots represent other cells. Red boxes indicate tumor FOVs, green boxes indicate normal liver FOVs, purple boxes represent both FOVs (containing both normal liver regions and malignant cell regions), and gray boxes indicate other FOVs.
